# A targeted folate receptor-α near-infrared fluorescent agent used for in vitro diagnosis of endometrial cytology

**DOI:** 10.1101/2020.02.23.20026948

**Authors:** Dongxin Liang, Xiaoqian Tuo, Lanbo Zhao, Kailu Zhang, Yiran Wang, Xue Feng, Panyue Yin, Lin Guo, Wei Jing, Qing Wang, Chao Sun, Junkai Zou, Lu Han, Qiling Li

## Abstract

The aim of this study is to perform the synthesis and evaluation of the new near**-**infrared targeting fluorescent dye folic acid-ZW800-1 (ZW-FA) and to explore its potential feasibility for in vitro diagnosis of endometrial cancer. Characterisation and Folate receptor-α (FR-α) targeting verification of ZW-FA were performed first and 92 patients were recruited, after liquid-based cytology preparations, during a 15-month period. ZW-FA and Hematoxylin-Eosin (H&E) staining were performed on all cytological slides successively; the histological diagnoses were regarded as the gold standard for ROC curve analysis. The cut-off value of ZW-FA fluorescence intensity is 62.9745; the sensitivity (Se), specificity (Sp), false-negative rate (FNR), false-positive rate (FPR), positive predictive value (PV+) percentage and negative predictive value (PV–) of the ZW-FA method are 84.6%, 85.2%, 15.4%,14.8%, 93.2% and 69.7%, respectively. ZW-FA is potentially efficient for in vitro diagnosis of endometrial lesions based on the FR-α expression level of different endometrial lesions.

## Introduction

Endometrial cancer is the second-most prevalent cancer after breast cancer, with a median diagnosed age of 62 years, and the five-year survival of early diagnosed cases is approximately 96% [1]. The limitation of dilation and curettage (D&C) has led to the advent of new and simple methods for endometrial sampling, including those by Pipelle, SAP-1 and Li Brush, together with the endometrial cytology test (ECT) [2-7]. However, limitations also exist in ECT for early diagnosis and screening of endometrial cancer, such as a lack of enough cytology pathologists and a unified standardisation system [8]. Immunocytochemical evaluation for phosphatase and tensin homolog deleted on chromosome 10 (PTEN), P53, Ki-67 and other markers has been tried previously; however, easier, cheaper and less time-consuming methods for early diagnosis and screening are still needed [9-11]..

Folate receptor-α (FR-α/FRA) is a 38- to 40-kDa molecule with a high affinity for folic acid and its derivatives, and a member of the folate receptors family, which consists of four known isoforms (including α, FRA; β, FRB; γ, FRG; and δ, FRD) [12, 13]. FR-α in normal tissues is restricted to apical surfaces of some organs such as the kidney, lung and choroid plexus [13]. Overexpression of FR-α has been reported in various solid tumours such as non–small cell endometrial cancer, cell lung adenocarcinoma, breast carcinoma, ovarian cancer and so on [14-17]. Based on the expression of FR-α, the agent used in intraoperative imaging, FRα-targeting antibody drugs and diagnosis of endometrial cancer, ovarian cancer and non–small cell lung cancer (NSCLC) were developed previously [14, 18-22]. Nevertheless, research regarding agents used in the diagnosis of endometrial cancer is rarely carried out yet.

Near-infrared (NIR) fluorescence imaging has emerged as a non-invasive, non-ionising and real-time visualisation technique, and compared to conventional fluorescent dyes, NIR dyes show ultralow autofluorescence, providing high signal-to-background ratio images [23]. Zwitterion NIR fluorophore (ZW800-1) was engineered for high hydrophilicity, no serum binding, ultralow non-specific tissue uptake, and rapid elimination from the body by renal filtration [24]. Based on these studies, a bridge between water-insoluble fluorescent dyes and live-cell fluorescence microscopy and protein targeting for cancer diagnosis and therapeutics is developing. Herein, we completed the combination of ZW800-1 and folic acid and constructed a new, near-infrared targeting fluorescent dye, folic acid-ZW800-1 (ZW-FA) (ZL201510104185.5). In this study, we performed the synthesis and evaluation of the new agent, ZW-FA, and aimed to explore its potential feasibility for in vitro diagnosis of endometrial cancer and precancerous lesions.

## Results

### Synthesis and characterisation of ZW-FA

The synthesis procedures of ZW-FA are presented in Figure 1A. As depicted in that figure and in the supplementary Table 1, NIR fluorophore ZW-FA was synthesised by employing p-hydrazinobenzenesulfonic acid and potassium hydroxide, together with a preparation of activated folic acid. The crude product was washed with the eluent consisting of acetone and water with a volume ratio of 1:3. NMR and mass spectrometry using electrospray ionisation (MS-ESI) were used to detect the target product during column chromatography. As implied in Figure 1B and 1C, the final ZW-FA product had the ability to emit near-infrared light.

**Figure 1.**
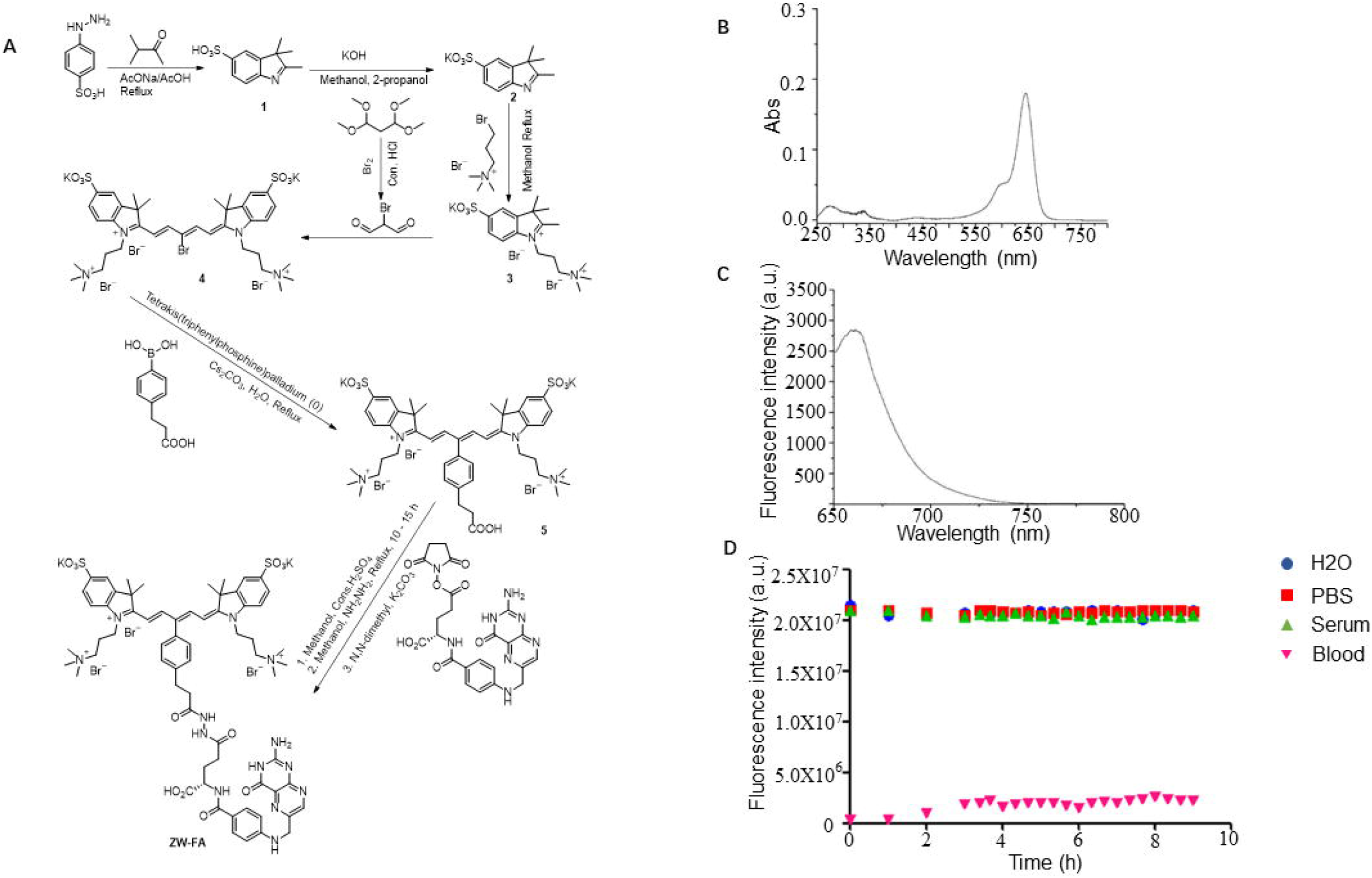
The synthesis and characterisation of ZW-FA. A Synthesis steps of ZW-FA. B Ultraviolet spectra of ZW-FA: the maximum absorption wavelength is around 645–650 nm. C Fluorescence spectra of ZW-FA: the emission wavelength is around 660 nm. D photostability of ZW-FA in water, PBS, serum and blood at 37□°C under continuous 650□nm laser exposure.

As shown in the synthesis procedures, using methanol as a solvent suggested the superior aqueous solubility of ZW-FA. We subsequently performed optical measurements at 37 °C in water, PBS, 100% FBS and blood. Excellent photostability of ZW-FA was observed by exposing the fluorescent dye in water, PBS, blood, and serum to a continuous irradiation with a 650□nm laser over long periods of study (∼8□h) (shown in Figure 1D).

### Uptake of ZW-FA and FR-α targeting verification

To evaluate the FR-α targeting efficiency and uptake of ZW-FA, we chose SKOV3, MDA-MB-231 (which overexpresses FR-α) and HUVEC (which expresses much less FR-α than SKOV3 does) cell lines to confirm the FR-α protein expression level further. As Patra CR et al. and Jones SK et al. reported previously, SKOV3 ovarian cancer cells and MDA-MB-231 breast cancer cells overexpress folate FR-α, and HUVEC cells express much lower levels of FR-α [25, 26]. Consistent findings were presented in this study (shown in Figure 2A–D), it was clearly observed that the intensity in the green channel, which corresponded to Human FOLR1 antibody fluorescence, was high in the case of SKOV3. Because ZW-FA was incubated following human FOLR1 antibody and Alexa Fluor 488, the human FOLR1 antibody first bound to the FR-α on the cell surface occupied by the folate receptor, which resulted in ZW-FA being blocked by cell uptake and binding; few FR-α were left that combined with ZW-FA (red colour zones shown by arrows in Figure 2A). We applied ZW-FA at the same concentration in MDA-MB-231, SKOV3 and HUVEC cells, and results showed that the intensity in the red channel was much higher in FR-α positive cell lines (SKOV3, MDA-MB-231) than in FR-α negative cell lines (shown in Figure 2E–H, 2I–L and 2M–P).

**Figure 2.**
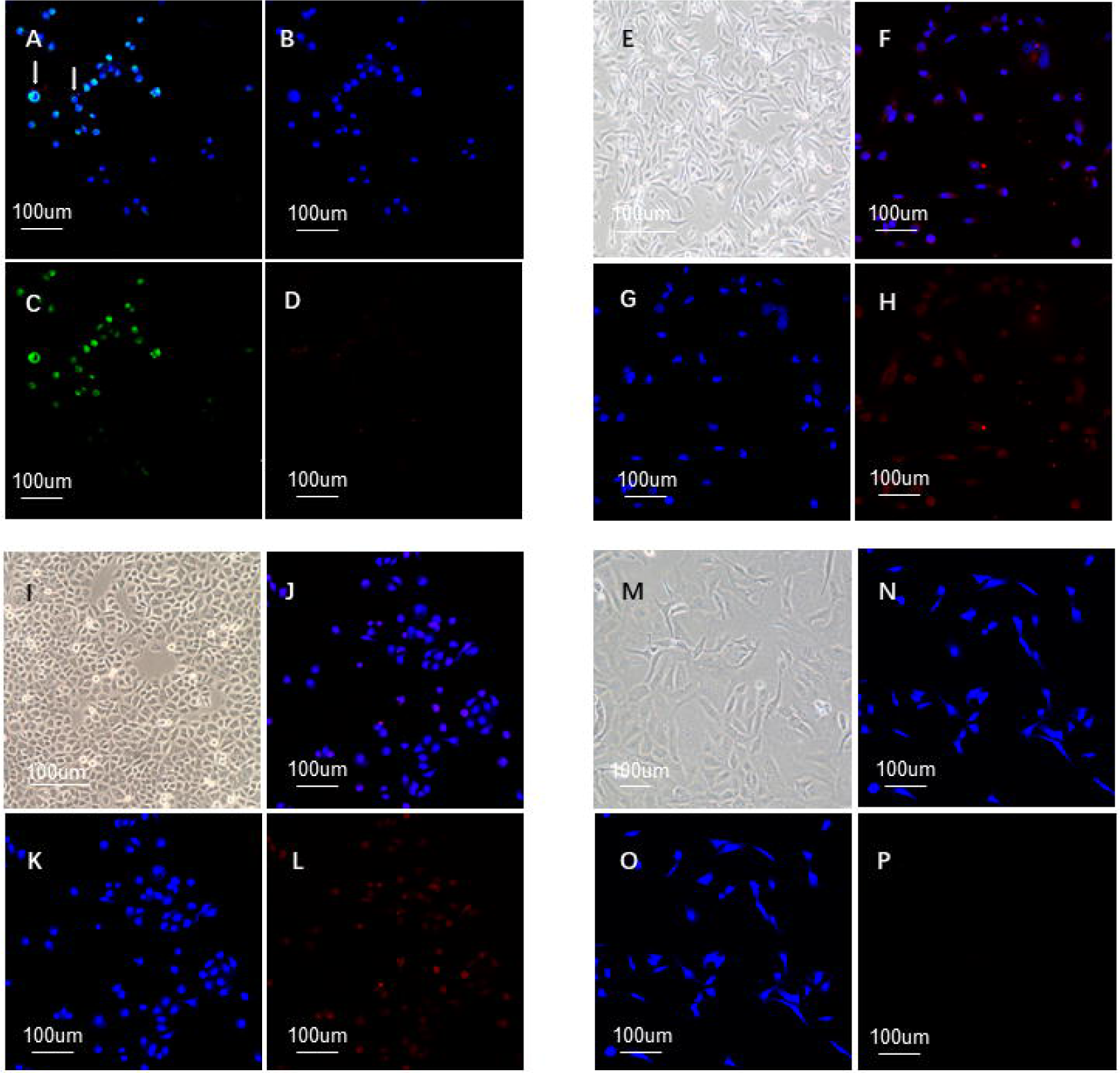
Fluorescence imaging of SKOV3, MDA-MB-231, HUVEC stained with human FOLR1 antibody and ZW-FA. A–D Fluorescence images of human ovarian cancer cell line SKOV3 (overexpressed FR-α) incubated with human FOLR1 antibody, ZW-FA and DAPI: (A) the montage image, (B) the blue channel to evaluate the nuclei stained with DAPI, (C) the green channel to evaluate the FR-α stained with human FOLR1 antibody, (D) the red channel to evaluate the FR-α stained with ZW-FA. E–L Fluorescence images of MDA-MB-231 and SKOV3 (overexpressed FR-α): (E, I) optical images of cell lines, (F, J) the montage image, (G, K) the blue channel to evaluate the nuclei stained with DAPI, (H, L) the red channel to evaluate the FR-α stained with ZW-FA. M–P Fluorescence images of HUVEC incubated with ZW-FA and DAPI: (M) optical images of cell lines, (N) the montage image, (O) the blue channel to evaluate the nuclei stained with DAPI, (P) the red channel to evaluate the FR-α stained with ZW-FA.

Because SKOV3 cells were incubated with ZW-FA in gradient concentration, the intensity in the red channel that corresponds to ZW-FA fluorescence significantly increased as concentration increased (shown in Figure 3A–C, 3D–F and 3G–I with gradient concentration separately).

**Figure 3.**
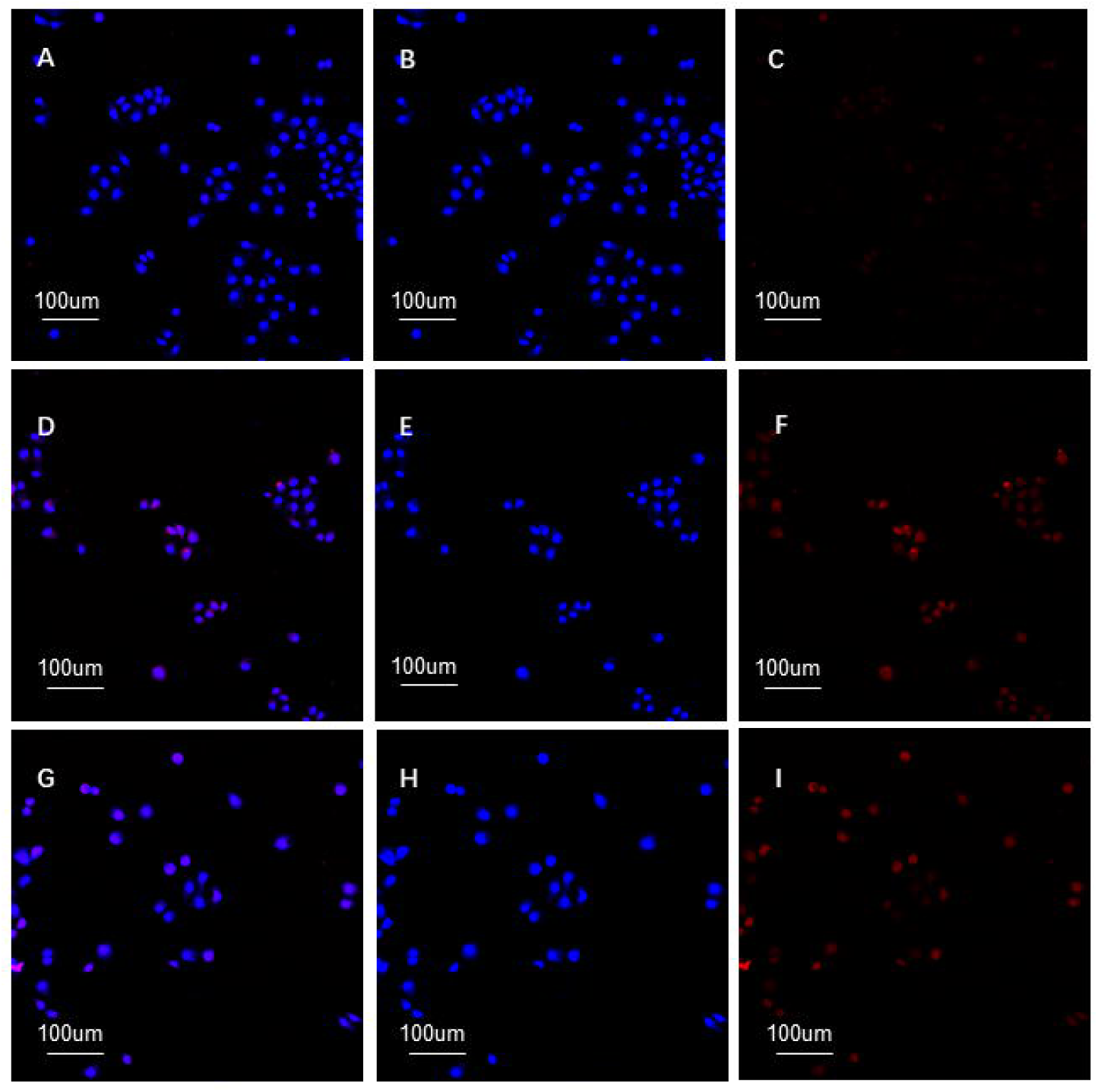
Fluorescence imaging of SKOV3 stained with ZW-FA in gradient concentration. A–I SKOV3 incubated with ZW-FA in 400, 500, 600 ug/ml separately: (A, D, G) the montage image, (B, E, H) the blue channel to evaluate the nuclei stained with DAPI, (C, F, I) the red channel to evaluate the FR-α stained with ZW-FA.

### Patients enrolled

Ninety-two patients were enrolled in this study. Among the histopathological diagnoses of endometrial samples, 61 were endometrial cancer, four were hyperplasia with atypia, eight were hyperplasia without atypia, five were atrophic endometrium, 11 were proliferative endometrium and three were secretory endometria. Detailed information of the histopathology and cytopathology of all samples is showed in the supplementary Table 2. Eight of 92 cytopathology slides were thought by pathologists to be unsatisfied samples; 55 of 65 samples divided in the Positive Group via histopathology were also diagnosed as positive, using ZW-FA, because significantly high fluorescence intensity was observed (shown in Figure 4A, B and C). The other 10 cases were misdiagnosed as negative, using ZW-FA for their relatively weak fluorescence intensity. Four of 27 samples (two hyperplasia without atypia and two proliferative endometrium) divided in the Negative Group via histopathology were misdiagnosed as positive, using ZW-FA. The rest of the samples had the same diagnoses, using these two methods (shown in Figure 4D, E and F). Interestingly, one case was diagnosed as endometrial cancer via D&C, hyperplasia with atypia via postoperative pathology, and endometrial cancer both via ZW-FA and cytopathology. Since cytology sampling was performed between D&C and operation; we regarded this case as endometrial cancer for ROC curve analysis.

**Figure 4.**
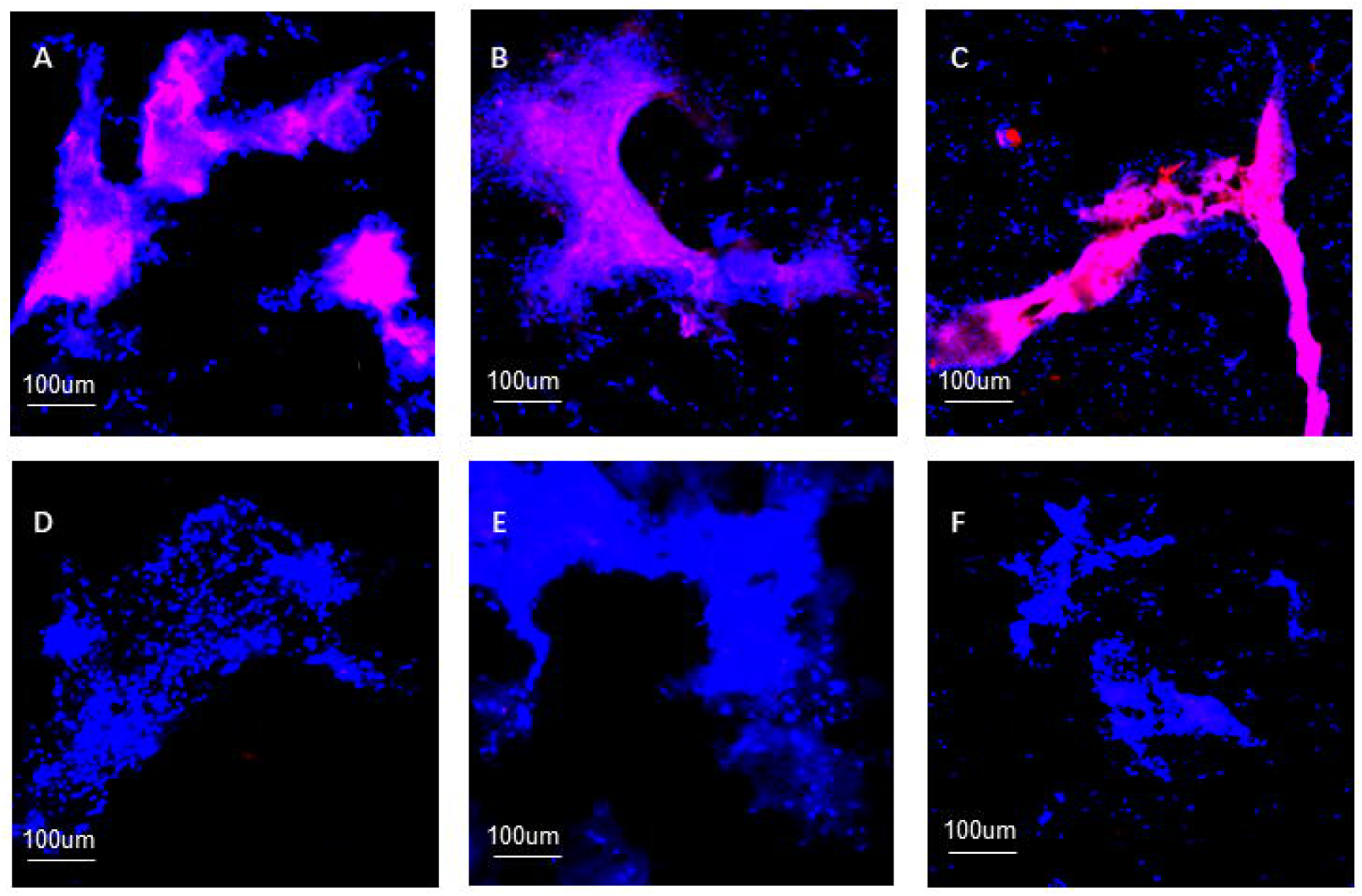
Fluorescence imaging of different endometrial lesions. A Endometrioid carcinoma. B clear-cell carcinoma. C atypical hyperplasia. D proliferative endometrium. E secretory endometrium. F atrophic endometrium.

### Diagnostic utility of ZW-FA staining

Because fluorescence intensity was obtained, in order to quantify fluorescence intensity, we herein introduced Image J to obtain the quantified intensity data. The ROC curve was then used to analyse further the quantified intensity data mentioned earlier. The area under the ROC curve (AUC) was 0.881 (shown in Figure 5), which indicated a highly accurate test, according to an arbitrary guideline (based on a suggestion by Swets) [27]. According to the Youden Index, 62.9745 was regarded as the cut-off value of ZW-FA fluorescence intensity, when Se was 84.6%, Sp was 85.2%, FNR was 15.4%, FPR was 14.8%, PV+ was 93.2% and PV– was 69.7%. In addition, the cytological diagnoses showed that 8 of 92 samples were regarded as unsatisfied samples, and 2 of 92 samples were misdiagnosed after two-step staining.

**Figure 5.**
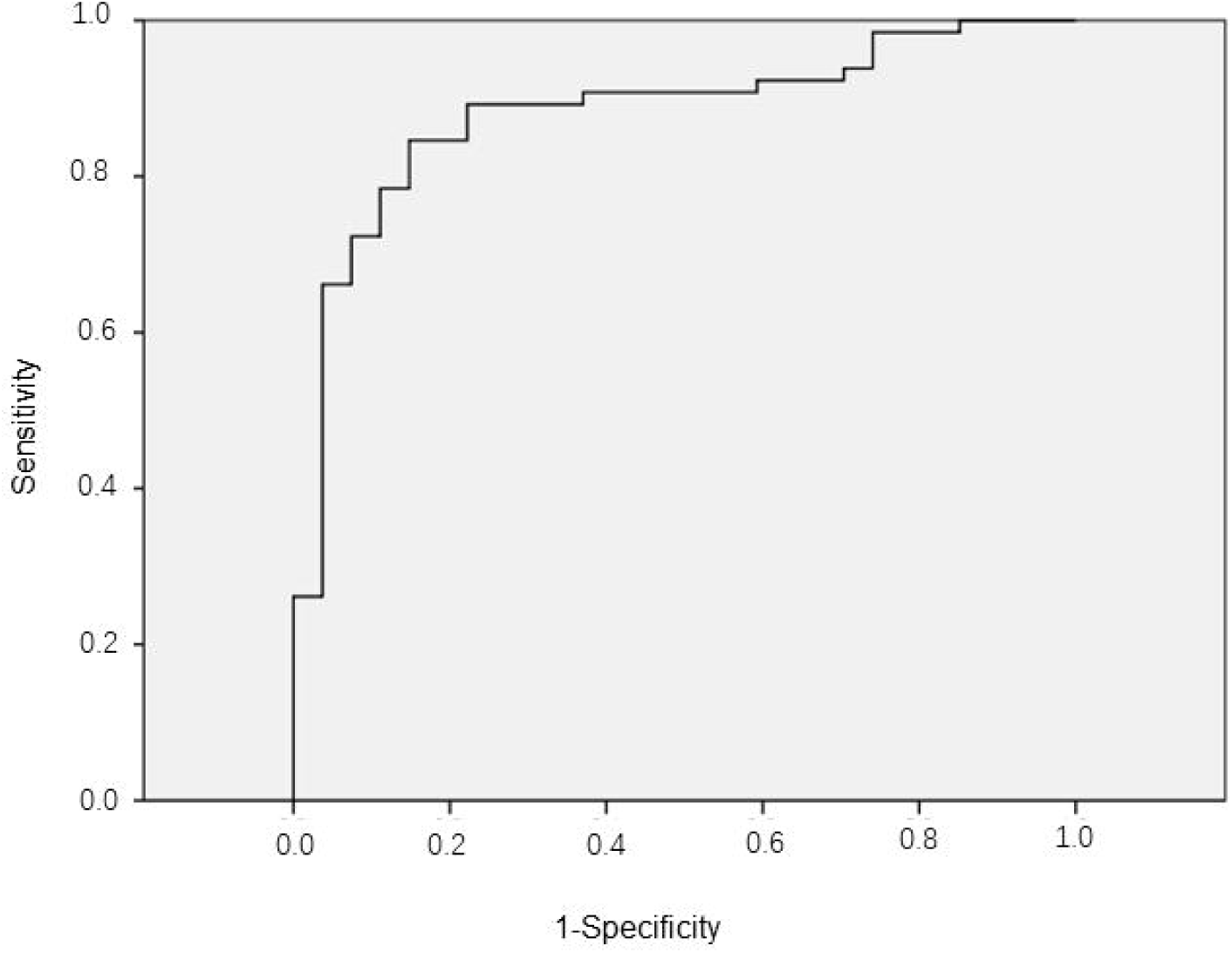
ROC curve. Area under the ROC curve (AUC) is 0.881, indicating a highly accurate test. n = 92.

## Discussion

The limitations of the ECT have promoted many studies to aid endometrial cytological diagnosing, such as immunocytochemical evaluation. However, easier, cheaper and less time-consuming methods for early diagnosis and screening are still needed [9, 11]. Because previous reports have indicated an increased expression of FR-α in a large number of patients with uterine cancer and other solid tumours, a potential strategy should be to distinguish benign, precancerous and malignant lesions based on the expression of FR-α [14, 17, 28, 29]. Consequently, the agent used in intraoperative imaging, FRα-targeting antibody-drugs and diagnosis of endometrial cancer, ovarian cancer and NSCLC were developed previously [14, 18-22]. Therefore, we synthesised the near-infrared targeting fluorescent dye ZW-FA, performed its characterisation and explored its potential feasibility in vitro diagnosis of endometrial cancer in this study.

We propose a potential method that uses ZW-FA to screen patients with endometrial lesions, based on its FR-α expression level. Potentially precancerous or malignant lesions will be further determined by histopathology or cytopathology, because the staining method used in this article rarely affects further cytological diagnoses.

In this study, four cases (two hyperplasia without atypia and two proliferative endometrium) were diagnosed by using ZW-FA as potentially precancerous or malignant lesions. As Senol et al. reported previously, 29.4% of endometrial hyperplasia was found with FR-α expression and 6.7% was found with high FR-α expression, and endometrial cancer was found with significantly high FR-α expression [30]. Consistently in this study, significantly higher FR-α expression was encountered in endometrial carcinomas in comparison to hyperplasia and normal endometrium.

Ten cases of endometrial cancer were diagnosed as potentially benign lesions via ZW-FA. Among these cases, one case was thought by pathologists to be an unsatisfied sample that might lead to misdiagnosis. As Brown Jones M et al. reported previously, the FR-α expression level was associated with histology subtypes, grade, advanced stage and disease□specific survival, which indicated that not all endometrial cancer expressed significantly high FR-α [31].

Of note, one case diagnosed as endometrial cancer by D&C and hyperplasia without atypia by postoperative histology, but a positive ZW-FA and cytopathology finding was reported in this study. A similar case was reported and meta-analysis was also performed previously, which prompted endometrial cytology as a method to improve the accuracy of diagnosis of endometrial cancer [32]. Herein, we inferred the potential accuracy of diagnosis of endometrial cancer by using ZW-FA staining.

Using ZW-FA for in vitro diagnosis of endometrial lesions is easier, cheaper and less time-consuming. Moreover, it could relieve the restriction of insufficient cytopathology specialists in rural areas if they could use cytopathology as the screening method, and using ZW-FA can primarily differentiate the overexpression type of endometrial lesions for further FR-α targeted therapy. Limitations also exist using this method, because not all types of endometrial lesions were included in this study, and the samples analysed in this research are still insufficient to support this method as a screening method in high-risk populations. Further validations are still needed. More data are needed before ZW-FA can be used for in vitro diagnosis as an effective screening tool for asymptomatic women with a high-risk of endometrial cancer.

The targeted folate receptor-α (FRA) near-infrared fluorescent agent ZW-FA is potentially efficient for in vitro diagnosis of endometrial lesions based on its FR-α expression level. The cut-off value of ZW-FA fluorescence intensity is 62.9745, and the Se, Sp, FNR, FPR, PV+ and PV– of the ZW-FA method are 84.6%, 85.2%, 15.4%, 14.8%, 93.2% and 69.7%, respectively. The diagnostic accuracy of endometrial carcinoma will be improved by subsequent histopathology or cytopathology. In a word, using ZW-FA for in vitro diagnosis as a potential screening method seems to be an easier, cheaper and less time-consuming method.

## Materials and Methods

### Synthesis of the ZW-FA compound

Synthesis of ZW-FA is composed of eight steps in all (shown in Figure 1A). The detailed steps and characterisation of each step are illustrated in the supplementary Table 1. In this part, 3-methyl-2-butanone and 4-hydrazinobenzenesulfonic acid were purchased from Aladdin; 4-(2-carboxyethyl) phenylboronic acid and 1,1,3,3-tetramethoxypropane were purchased from Alfa Aesar; and 3-Bromo-N,N,N-trimethylpropan-1-aminium bromide was purchased from Ark Pharm. All solvents and other reagents were of reagent grade quality and purchased commercially.

### Characterisation

^1^H and ^13^C NMR spectra were recorded on a Varian unity INOVA-400 spectrometer at 400, using TMS as an internal standard for ^1^H NMR spectra. Mass spectra were performed with a Bruker micrOTOF-Q II ESI-Q-TOF LC/MS/MS spectrometer. Fluorescence spectra were recorded by a Hitachi F-4500 (Tokyo, Japan) instrument. Photostability of ZW-FA was evaluated in a variety of biological media, including water, PBS, serum and blood at 37□°C under continuous 650□nm laser exposure (Cary series UV-Vis; Agilent Technologies, CA, USA).

### Uptake of ZW-FA and FR-α targeting verification

The FR-α positive cell line human ovarian cancer SKOV3 and human breast cancer MDA-MB-231 were obtained from the Shanghai Cell Bank of the Chinese Academy of Sciences (Shanghai, China), and the FR-α negative cell line HUVEC was obtained from ATCC (USA). The SKOV3 and HUVEC were grown in folate-free RPMI 1640 medium (Invitrogen, IL) and supplemented with 10% fetal bovine serum (FBS), 10 U/mL penicillin and 10 mg/mL streptomycin. Cultures were maintained at 37 °C under humidified conditions with 5% CO_2_. To compare the uptake and verify the FR-α targeting efficiency in SKOV3 and HUVEC, cells were cultured on coverslips in a 24-well plate at 10^5^/well overnight. Each well was washed three times with PBS, treated with 0.5 mL of 4% paraformaldehyde for 30 min to fix the cells and then washed with PBS. They were then incubated in ZW-FA only or human FOLR1 antibody (R&D, USA, catalogue #MAB5646) overnight and followed by ZW-FA for one hour, washed by PBS again and incubated with DAPI (Roche, Switzerland, catalogue #10236276011) for 30 minutes. Cells incubated with human FOLR1 antibody then needed to be incubated with Alexa Fluor 488 (Abcam, Australia, catalogue #ab150117) later. Finally, the antifade mounting medium (Beyotime, China, catalogue #P0126) was used for sealing, and the fluorescent inverted microscope (Nikon Eclipse Ti, Japan) and laser confocal microscope (C2, Nikon, Japan) were used to take photos. The excitation wavelengths of 408 nm, 488 nm and 633 nm were used to assess the fluorescence of DAPI, human FOLR1 antibody and ZW-FA, respectively. Of note, the steps of incubation and afterwards needed to be protected from light.

### Patients and sample collection

This study was approved by the Ethics Committee of the First Affiliated Hospital of Xi’an Jiaotong University, and informed written consent was obtained from all subjects before this study. In all, 92 patients who underwent total hysterectomies or D&Cs were enrolled in this study during a 15-months period (07/2018 to 10/2019). The patients’ final diagnoses were confirmed by postoperative histological examination. The histological diagnostic types, according to the International Society of Gynecological Pathology Classification, included endometrial carcinoma, hyperplasia with atypia, hyperplasia without atypia, atrophic endometrium, proliferative endometrium and secretory endometrium. The cytological diagnosis was made according to the criteria formulated by Fox, ranging from benign cells (including atrophic, proliferative and secretory endometrium), non-atypical hyperplastic cells and atypical hyperplastic cells to malignant cells. The diagnoses of endometrial carcinoma and hyperplasia with atypia were grouped into the Positive Group and, vice versa, others were grouped into the Negative Group.

### Cytological preparation

Endometrial cytological samples were all collected from patients by using the Li Brush (20152660054, Xi’an Meijiajia Medical Technology Co. Ltd., China). The following procedures were performed as described in a previous study [8, 33-35].

### Two-step staining for endometrial slides

First, the clips were taken out of the alcohol and washed for five minutes three times by phosphate-buffered saline (PBS). The clips of endometrial cells were incubated with ZW-FA (500 ug/ml) for two hours and washed with PBS five minutes three times again. The clips were then incubated with DAPI (CST) for one hour (h). Finally, they were washed three times and prepared for imaging. They were examined immediately using an appropriate excitation wavelength. After interpreting the slides, H&E staining was performed as described in a previous study [8], and cytological diagnoses were finally processed.

### Data analysis and statistics

After performing ZW-FA staining, the slides were evaluated and typical photos were later taken using the laser confocal microscope. We used the excitation wavelength of 408 nm to assess DAPI fluorescence, and the excitation wavelength of 633 nm to assess ZW-FA fluorescence. The cytological and histopathological results of all slides were evaluated by two experienced pathologists, using conventional optical microscopy. The fluorescence in the photos of every slide was analysed using Image J, and finally we got the intensity data of all slides. To obtain a cut-off value of the intensity data to differentiate endometrial cancer and precancerous endometrial lesions from benign endometrium, we introduced a receiver operating characteristics (ROC) curve to analyse. Comparison of ZW-FA and histopathology was performed subsequently to calculate sensitivity (Se), specificity (Sp), false-negative rate (FNR), false-positive rate (FPR), positive predictive value (PV+) and negative predictive value (PV–).

## Data Availability

All the data supporting the findings of this study are available within the article and its supplementary information files and from the corresponding author upon reasonable request.

## Acknowledgements

We thank Professor Wu Biao and Dr. Yang Dong of Northwest University for synthesizing ZW-FA. This work was supported by the Major Basic Research Project of Natural Science of Shaanxi Provincial Science and Technology Department (2018JM7073, 2017ZDJC-11), the Clinical Research Award of the First Affiliated Hospital of Xi’an Jiaotong University, China (XJTU1AF-2018-017, XJTU1AF-CRF-2019-002), the Key Research and Development Project of Shaanxi Provincial Science and Technology Department (2017ZDXM-SF-068, 2019QYPY-138), the Shaanxi Provincial Collaborative Technology Innovation Project (2017XT-026, 2018XT-002), and the Medical Research Project of Xi’an Social Development Guidance Plan (2017117SF/YX011-3). The funders had no role in study design, data collection and analysis, decision to publish, or preparation of the manuscript.

## Author contributions

Qiling Li designed and configured the study. Dongxin Liang and Xiaoqian Tuo conducted the experiments and wrote the paper. Lanbo Zhao, Qing Wang, Chao Sun, Junkai Zou helped to conduct the experiments. Kailu Zhang, Yiran Wang, Xue Feng, Panyue Yin, Lin Guo, Wei Jing and Lu Han collected the samples. Lanbo Zhao helped to polish the manuscript. Dongxin Liang and Xiaoqian Tuo contributed equally to this work.

